# Restoration of leukomonocyte counts is associated with viral clearance in COVID-19 hospitalized patients

**DOI:** 10.1101/2020.03.03.20030437

**Authors:** Xiaoping Chen, Jiaxin Ling, Pingzheng Mo, Yongxi Zhang, Qunqun Jiang, Zhiyong Ma, Qian Cao, Wenjia Hu, Shi Zou, Liangjun Chen, Lei Yao, Mingqi Luo, Tielong Chen, Liping Deng, Ke Liang, Shihui Song, Rongrong Yang, Ruiying Zheng, Shicheng Gao, Xien Gui, Hengning Ke, Wei Hou, Åke Lundkvist, Yong Xiong

## Abstract

**Background:** Viral clearance is one important indicator for the recovery of SARS-CoV-2 infected patients. Previous studies have pointed out that suboptimal T and B cell responses can delay viral clearance in MERS-CoV and SARS-CoV infected patients. The role of leukomonocytes in viral clearance of COVID-19 patients is not yet well defined.

**Methods:** From January 26 to February 28, 2020, an observational study was launched at the Department of Infectious Diseases, Zhongnan Hospital of Wuhan University, Wuhan, China. We enrolled 25 laboratory-confirmed COVID-19 patients, whose throat-swab specimens were tested positive for SARS-CoV-2 infection by qRT-PCR. To investigate the factors that contribute to the viral clearance, we comprehensively analyzed clinical records, counts of lymphocyte subsets including CD3+, CD4+, CD8+ T cells, B cells and NK cells in the patients who successfully cleared SARS-CoV-2, and compared to those that failed to, after a standardized treatment of 8-14 days.

**Findings:** In 25 enrolled COVID-19 patients, lymphopenia was a common feature. After the treatment, 14 out of the 25 enrolled patients were tested negative for SARS-CoV-2. The patients that cleared the infection had restored the numbers of CD3+, CD4+, CD8+ T cells and B cells as compared to the still viral RNA positive patients, while the recovered patients had a higher count of leukomonocytes.

**Conclusions:** By comparison of leukomonocytes counts in COVID-19 patients at different stages of the disease, we found that CD3+, CD4+, CD8+ T cells and B cells appear to play important roles in viral clearance. The restoration of leukomonocytes counts from peripheral blood can be used as prognosis for the recovery of an COVID-19 infection. We propose that restoration of leukomonocytes counts can be added to the COVID-19 diagnostic guidance as a criterion for releasing and discharging patients.

## Introduction

In December 2019, a cluster of novel coronavirus-associated pneumonia cases was emerging in Wuhan, China [1]. The disease rapidly spread from Wuhan and caused a wide epidemic in China. The World Health Organization (WHO) has recently named the disease as Coronavirus Disease-2019 (COVID-19) [2]. The etiology of the disease is a novel β-coronavirus, taxonomically named as severe acute respiratory syndrome coronavirus 2 (SARS-CoV-2). Until February 29, 2020, SARS-CoV-2 causes an epidemic worldwide. More than 80,000 people in over 60 countries or territories have been contracted of COVID-19, and more than 3000 people have died [3].

The clinical manifestations of COVID-19 cases range from an asymptomatic course to serve acute respiratory symptoms and even acute respiratory failure [4-7]. Transmission occurs from human to human with nosocomial transmissions. The incubation time of COVID-19 has been reported from 0 to 24 days and also asymptomatic carriers can be infectious [8].

As a zoonotic virus, the natural host of SARS-CoV-2 is believed to be bats [9], however, the process of cross-species transmission(s) is still not known. The very first cases were associated with the Huanan Seafood Wholesale Market in Wuhan, China [10]. There is no effective antiviral drug available yet. Although the pathogenesis of COVID-19 is still unclear, lymphopenia has been found in most COVID-19 patients [10-12]. This feature has also been found in the other two fatal human coronavirus infections caused by severe acute respiratory syndrome coronavirus (SARS-CoV) and the Middle East respiratory syndrome coronavirus (MERS-CoV) [13, 14]. Previous studies showed that T cells, especially CD4+ and CD8+ T cells, play a significant antiviral role in balancing the combat against MERS-CoV or SARS-CoV infections [15]. These results suggested that leukomonocytes counts could be critical indicators associated with severity and the disease outcome. In this study, we retrospectively analyzed 25 COVID-19 cases to elucidate the dynamic of lymphocyte subset changes in the peripheral blood and their roles during the viral clearance.

## Methods

### Study design

From January 26 to February 28, 2020, 25 COVID-19 patients who had been repeatedly analyzed for lymphocyte subsets and SARS-CoV-2 RNA were enrolled in the study. Informed consents were obtained from all patients upon admission to the Department of Infectious Diseases, Zhongnan Hospital of Wuhan University, Wuhan, China.

### Data collection

The medical records, including epidemiological and demographic information, clinical manifestations, laboratory data, and outcome of disease, were collected. To avoid subjective biases, two investigators reviewed the data independently and made a final consensus.

Peripheral blood (100 μl) was collected from patients at admission to hospital and after 7 – 10 days of standard therapy (Version 6, 2020 Feb 18, http://www.nhc.gov.cn/yzygj/s7653p/202002/8334a8326dd94d329df351d7da8aefc2.shtml). Lymphocyte subsets such as CD3+, CD4+, CD8+ T cells, B cells and NK cells were stained by using the BD Multitest™IMK kit (BD Ltd., San Jose, CA, USA) according to the manufacturer’s instruction. The lymphocyte subset analyses were performed by flow cytometry (BD FACSCanto™II Flow Cytometer).

Serial throat-swab samples were taken from patients at the admission to hospital during 8 – 14 days of standardized treatment (see Table 1) according to the recommendation in the guideline. The swab samples were placed in 150 μL of virus preservation solution and viral RNA subsequently extracted as described earlier [5, 16]. Two target genes of the SARS-CoV-2 genome, including the open reading frame 1ab (ORF1ab) and the nucleocapsid protein (N) gene, were simultaneously amplified and tested in the real-time RT-PCR assay by using a SARS-CoV-2 nucleic acid detection kit according to the manufacturer’s protocol (Shanghai Bio-Germ Medical Technology Co Ltd, Shanghai, China). The reaction mixture consisted of 12 μL reaction buffer, 4 μL of enzyme solution, 3 μL of diethyl pyrocarbonate–treated water, 2 μL of RNA template, and 4 μL of probe/primers solution, which contained two sets of probe primers: one set targets towards to ORF1ab (forward prime 5’-CCCTGTGGGTTTTACACTTAA-3’; reverse primer 5’-ACGATTGTGCATCAGCTGA-3’; and the probe 5’-VIC-CCGTCTGCGGTATGTGGAAAGGTTATGG-BHQ1-3’), and another targets towards to N (forward primer 5’-GGGGAACTTCTCCTGCTAGAAT-3’; reverse primer 5’-CAGACATTTTGCTCTCAAGCTG-3’; and the probe 5’-FAM-TTGCTGCTGCTTGACAGATT-TAMRA-3’).

**Table 1.**
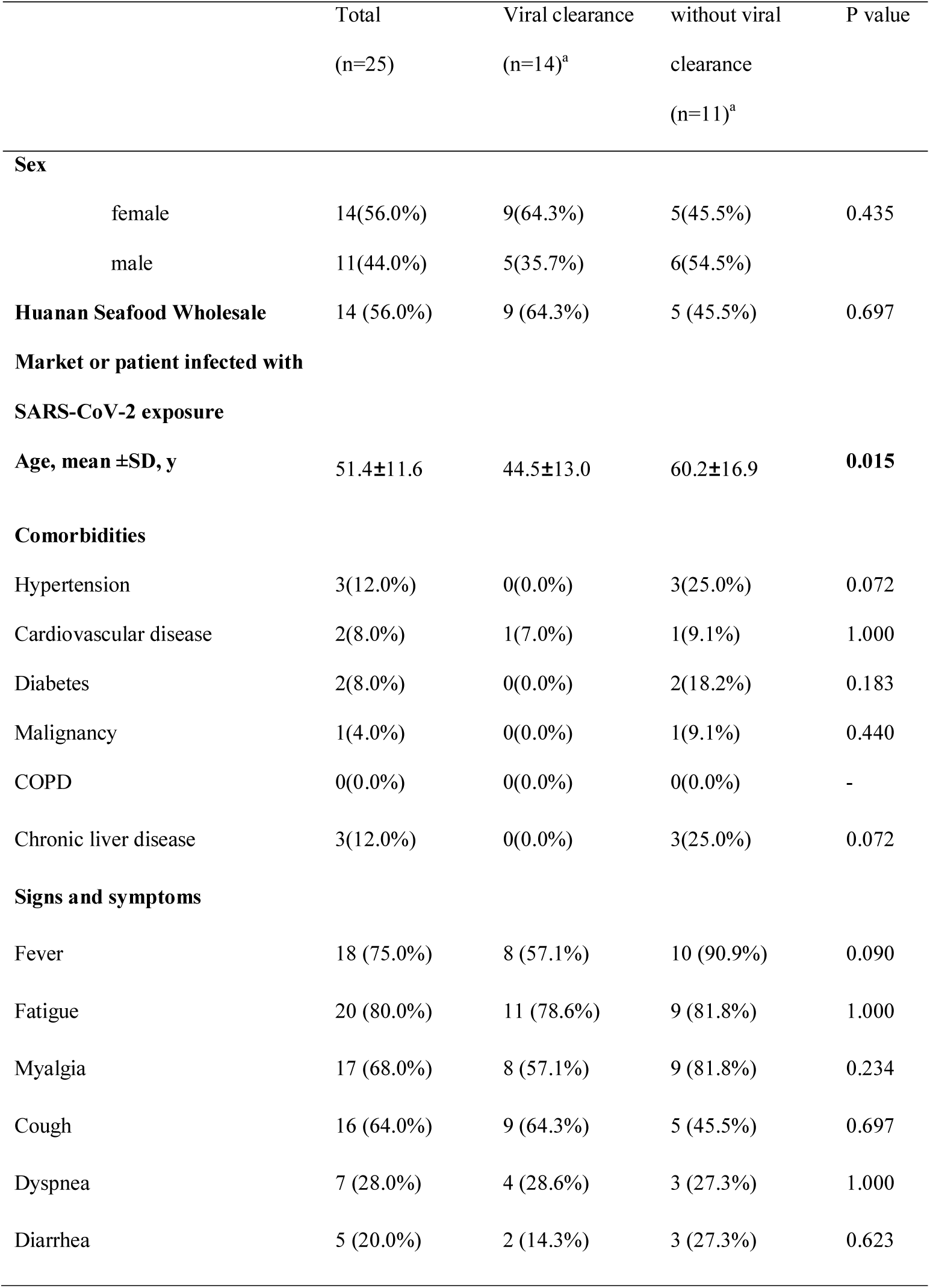
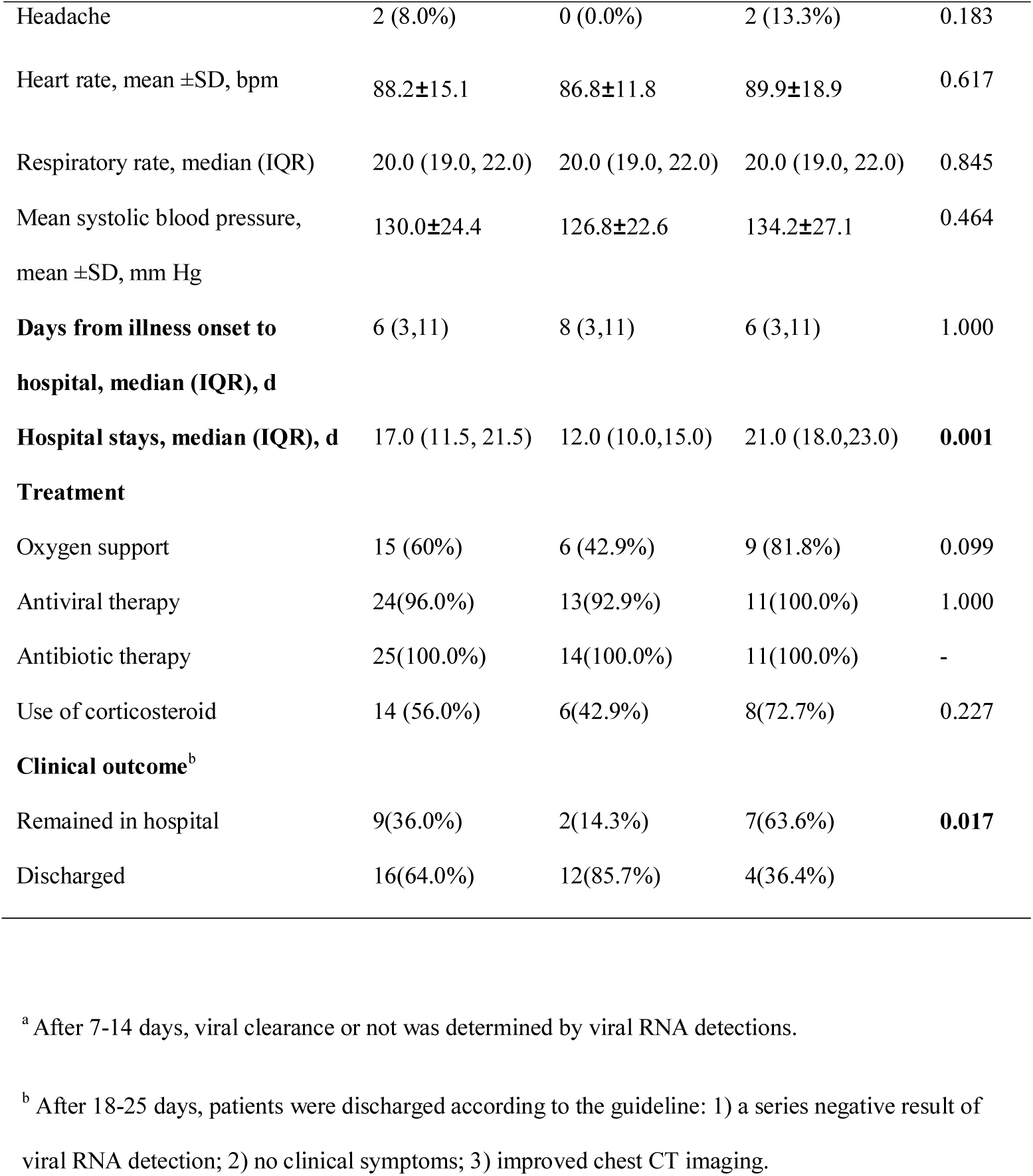
Demographics, baseline characteristics, treatment, and clinical outcomes of 25 patients infected with SARS-CoV-2

The results of the real-time RT-PCR analyzes were interpreted as: a cycle threshold value (Ct-value) less than 37 was defined as a positive result; a Ct-value of 40 or more was defined as a negative result; a medium load result, defined as a Ct-value of 37 to less than 40, required another test for confirmation. The patients who repeatedly tested SARS-CoV-2 RNA negative for at least two times with an interval of more than 1 day were regarded as viral negative. These diagnostic criteria were based on the recommendation by the National Institute for Viral Disease Control and Prevention, China (http://ivdc.chinacdc.cn/gjhz/jldt/202002/P020200209712430623296.pdf).

### Statistical analysis

We used the SPSS 17.0 software package for the statistical analyses. We used X^2^ tests or Fisher’s exact tests for categorical variables. For measurement of the data, a normal distribution was tested first and then proceeded to t-test, expressed as the mean ± standard deviations; otherwise the Mann-Whitney U test was used for non-normal distribution data, expressed in terms of median (25%–75% interquartile range, IQR). A p value of < 0.05 was considered statistically significant.

### The principle of medical ethics

This study was approved by the ethics board in Zhongnan Hospital of Wuhan University, Wuhan, China (No.2020011).

## Results

### Baseline Characteristics of COVID-19 patients

A total of 25 patients, 11 men and 14 women were enrolled in this study (Table 1). The ages of the patients spanned from 25 to 80 years (average 51.4±16.6 years). Fourteen people described a previous exposure history to the source of infection, either from the Huanan Seafood Wholesale Market or through direct contacts with COVID-19 patients. The most common symptoms at the onset of illness were: fatigue (80.0%), fever (37.4–39.1°C, 75.0%), myalgia (68.0%), cough (64.0%), and less common: dyspnea (28.0%) and diarrhea (20.0%). Seven cases had underlying diseases such as hypertension, cardiovascular disease, diabetes, malignancy or chronic liver disease (Table 1).

### Laboratory analysis before and after treatment

Laboratory analysis were performed before the treatment. The biochemical tests included alanine aminotransferase, aspartate aminotransferase, aspartate aminotransferase, creatinine, and D-dimer, which all were found normal (Table 2). However, blood counts of the patients showed leucopenia (4.5±1.9 ×10^9^/L) and lymphopenia (< 1.3 ×10^9^/L). The counts of CD3+ T cells, CD4+ T cells, CD8+ T cells, B cells, and NK cells were found lower than normal values. Interleukin-6 had increased to a mean of 16.1 (3.7 – 31.4) pg/ml. Five patients had an increase of procalcitonin (≥0.05 ng/mL) (Table 2).

**Table 2.** Laboratory results of 25 patients with the outcome of viral clearance.

|  | Normal<br>Range | Total | Viral clearance(n=14) | without viral<br>clearance (n=11) | P value |
| --- | --- | --- | --- | --- | --- |
| White blood cell Count ($\times 10^9$ /L) | 3.5-9.5 | 4.5±1.9 | 4.5±1.7 | 4.5±2.2 | 0.919 |
| Lymphocyte count ($\times 10^9$ /L) | 1.1-3.2 | **0.9 (0.6, 1.3)** | 0.8 (0.6, 1.4) | 0.9 (0.6, 1.2) | 0.565 |
| Monocyte count ($\times 10^9$ /L) | 0.1-0.6 | 0.37±0.15 | 0.33±0.15 | 0.42±0.14 | 0.117 |
| Neutrophil count ($\times 10^9$ /L) | 1.8-6.3 | 2.8 (1.5, 4.5) | 3.1 (1.6, 4.8) | 2.6 (1.3, 4.3) | 0.805 |
| Platelet count ($\times 10^9$ /L) | 125-350 | 179.5±68.8 | 191.5±70.4 | 164.3±66.7 | 0.336 |
| Alanine aminotransferase (U/L) | 9-50 | 24 (20.5, 38.5) | 21 (19, 50.5) | 25 (21.0, 35.0) | 0.583 |
| Aspartate aminotransferase (U/L) | 15-40 | 26 (21.0, 33.0) | 23.0(17.0, 34.8) | 28.0(26.0,33.0) | 0.066 |
| Total bilirubin ( mmol/L) | 5-21 | 9.6 (8.0, 12.7) | 9.6 (7.6, 12.2) | 10.7 (8.2, 16.7) | 0.511 |
| Lactate dehydrogenase (U/L) | 125-243 | 197(164.5, 286.5) | 197.0(155.0, 340.0) | 201.0(174.0, 279.0) | 0.956 |
| Prothrombin time (s) | 9.4-12.5 | 12.3 (11.7, 13.0) | 12.6 (11.7,13.5) | 12.3 (12.1,12.4) | 0.641 |
| Activated partial thromboplastin time (s) | 25.1-36.5 | 29.9±3.0 | 29.8±2.6 | 30.0±3.6 | 0.860 |
| D-dimer, (mg/L) | 0-500 | 187.0 (125.0,309.5) | 181.0 (104.5,271.0) | 233.0 (145.0,630.0) | 0.179 |
| Creatinine (μmol/L) | 64-104 | 65.5±14.6 | 64.3±14.3 | 67.0±15.6 | 0.659 |
| Procalcitonin, ng/mL ≥0.05, No. (%) | <0.05 | **5 (20.0%)** | 3 (21.4%) | 2 (18.2%) | 1.000 |
| Interleukin-6 (pg/ml) | 0-7 | **16.1 (3.7, 31.4)** | 13.8 (5.3, 30.2) | 17.6 (2.8, 49.9) | 0.805 |
| CD3+ T cell count ($\times 10^9$ /L) | 805-4459 | **561.0 (338.0, 770.0)** | 639.0 (400.3, 1038.0) | 485.0 (384.0, 732.0) | 0.311 |
| CD4+ T cell count ($\times 10^9$ /L) | 345-2350 | **297.0 (171.5, 447.5)** | 327.0 (182.8, 560.5) | 220.0 (148.0, 420.0) | 0.262 |
| CD8+ T cell count ($\times 10^9$ /L) | 345-2350 | **272.9±129.7** | 291.4±149.3 | 249.5±101.6 | 0.435 |
| B cell count ($\times 10^9$ /L) | 240-1317 | **111.5±75.8** | 133.2±84.8 | 83.8±54.2 | 0.107 |
| NK cell count ($\times 10^9$ /L) | 210-1514 | **170.0 (67.5, 245.0)** | 193.5 (75.0, 286.0) | 170.0 (43.0, 215.0) | 0.511 |

The treatment was mainly of supportive care (Table 1). Twenty-four patients were given antiviral therapy including arbidol (orally, 200 mg, three times per day), and oxygen support. Antibiotic therapy, both orally and intravenous, were given as described in Table 1. Fourteen patients received corticosteroids to suppress an excessive inflammatory activation. After standardized treatment of 8–14 days, according to the laboratory analysis of viral RNA, fourteen patients showed viral clearance and 11 patients were still positive for SARS-CoV-2 viral RNA (Table 1).

When comparing the group that showed viral clearance and the group that failed, we found no significant association to sex, epidemiological exposure history, comorbidities, onsets of signs and symptoms, or treatment, but with the age and days of hospitalization. The mean days of hospitalization in the group that showed viral clearance were 12 days (10.0 – 15.0), which was shorter as compared to the group that failed to clear the virus, for which the average days were 21 (18.0 – 23.0) days. Age can play an important role during the viral clearance: patients without viral clearance were older (60.2±16.9, y) than patients with viral clearance (44.5 ± 13.0 y), showing a statistically significant difference (Table 1).

### Association between peripheral lymphocyte counts and disease outcomes

Since lymphopenia was found in all the patients in the current study and also earlier reported elsewhere [10], we further analyzed the association between peripheral lymphocyte counts and disease outcomes. In the 14 patients with viral clearance after the 8 – 14 days of treatment, the counts of CD3+ T cells, CD4+ T cells, CD8+ T cells, and B cells were restored close to normal levels and with a significant difference as compared to the counts at admission in 12 patients. In the remaining 2 patients with viral clearance the leukomonocytes counts were still at low levels (Figure 1, A-E). However, in the patients without viral clearance (No.=11), no differences were seen before and after treatment regarding the CD3+, CD4+, CD8+, B, and NK cell counts (Figure 1, F-J). The association between peripheral lymphocyte counts and viral detection suggested that higher counts of CD3+ T cells, CD4+ T cells, CD8+ T cells, and B cells had a significant impact on the viral clearance.

**Figure 1.**
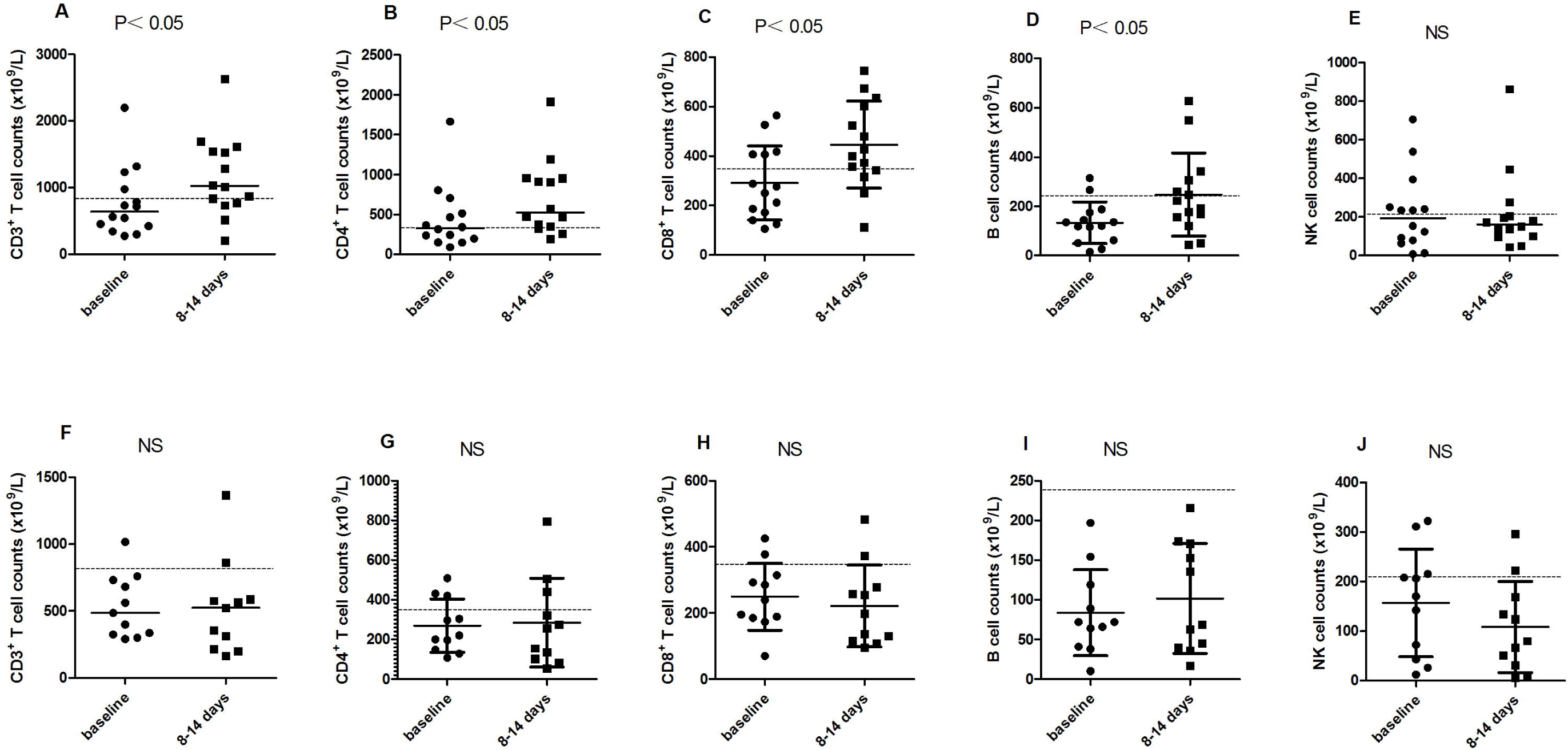
Comparison of peripheral changes in CD3+, CD4+, CD8+ T, B, and NK cell counts in patients who were negative for the SARS-CoV-2 (A-E) and positive (F-J) after 8-14 days of treatment. The medium number of peripheral lymphocyte counts are represented in the solid line. The threshold of normal peripheral lymphocyte counts is represented in the dash line. NS means no significant difference tested in statistical analysis: Mann-Whitney U test was used for A, B, E, F since these data are non-normal distributed and t-test was used for the other normal distributed data).

To further monitor the disease outcome of those 11 patients who were still positive for SARS-CoV-2 virus RNA, we extended the same treatment to 15 – 23 days (Figure 2). At 15 –23 days, 7 patients were negative for viral RNA detection (data for leukomonocyte subsets was available for 6 of these 7 patients), but 4 patients were still positive (all these four had been tested for leukomonocyte subsets). The 4 viral RNA positive patients had low counts of CD3+ T cells (< 1200 ×10^9^/L), CD4+ T cells (< 700 ×10^9^/L), CD8+ T cells (< 500 ×10^9^/L), B cells (< 400 ×10^9^/L), and NK cells (< 400 ×10^9^/L). Due to the small sample size, no statistically significant differences for the cell counts were observed, except for the NK cell counts, which showed a significant decrease in the 4 viral RNA positive patients (Figure 2, J). However, we found a similar trend that the increased counts of CD3+ T cells and CD8+ T cells corresponded to cases which were negative for viral RNA detection (5 out of 6). One case that still had low counts of CD3+ T cells, CD4+ T cells, CD8+ T cells, B cells and NK cells was found negative for viral RNA.

**Figure 2.**
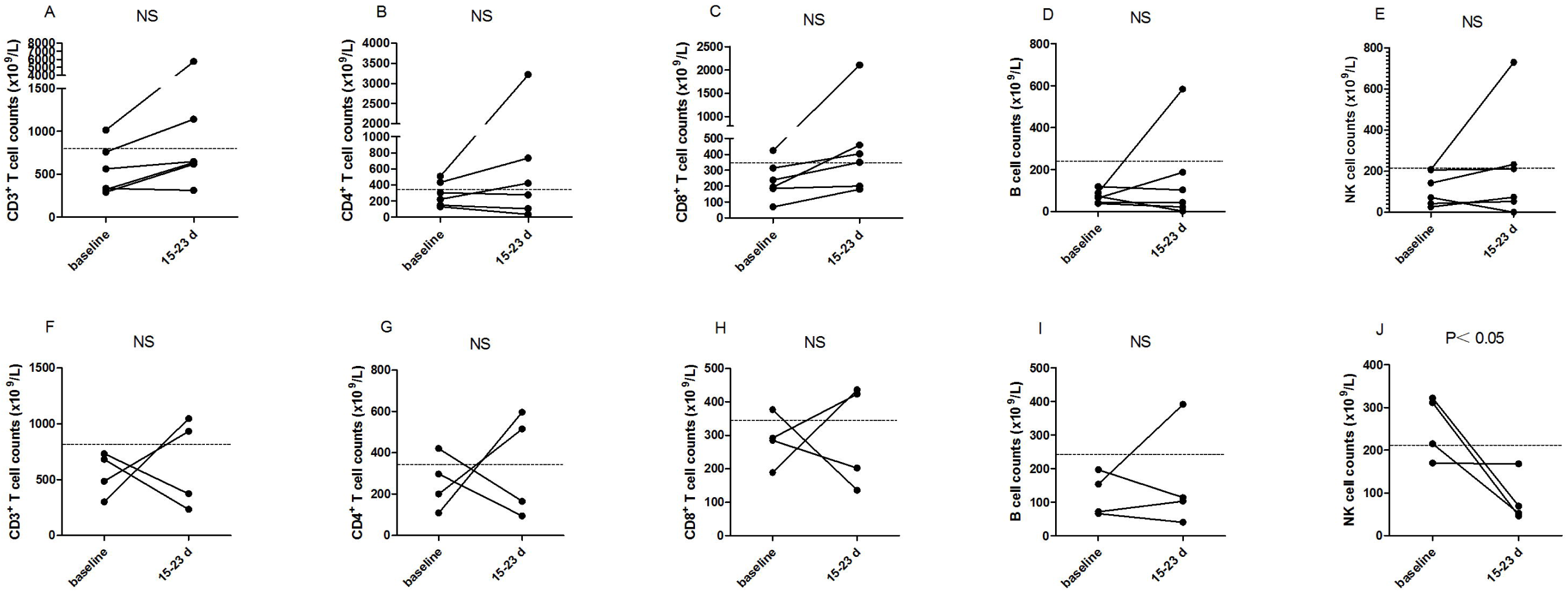
Comparison of peripheral changes in CD3+, CD4+, CD8+ T, B, and NK cell counts in six patients who were negative for the SARS-CoV-2 (A-E) and four patients who were positive (F-J) after 15-23 days of treatment. The medium number of peripheral lymphocyte counts are represented in the solid line. The threshold of normal peripheral lymphocyte counts is represented in the dash line. NS means no significant difference tested in statistical analysis and all data used Mann-Whitney U test.

### Clinical outcome

We observed the clinical course of 25 COVID-19 patients within 23 days of treatment. During the first phase of treatment, at 8 – 14 days, fourteen patients showed viral clearance and 12 of them were discharged from the hospital according to the guideline. The treatment of oxygen support therapy and clinical observation was continued for the remaining 2 patients in this group. Eleven viral RNA positive patients received a second phase of treatment between 15– 23 days. Until February 28, 7 of these 11 patients turned out to be viral RNA negative. Four of these 7 patients were then discharged from the hospital, while the other 3 (one had a liver disease and the other two still had significant clinical symptoms), together with the 4 patients who were still positive for viral RNA, were kept under treatment (Table 1). In total, 9 out of the 25 COVID-19 patients were still hospitalized after 23 days of clinical treatment.

## DISCUSSION

In this report, we retrospectively investigated the changes of CD3+, CD4+, CD8+, B, and NK cell counts in peripheral blood of 25 COVID-19 patients during the viral infection and the mechanisms for viral clearance. We found that restoration of CD3+, CD4+, CD8+ T cells, B cells and NK cells was associated the viral clearance in COVID-19 patients.

According to the Guideline of the treatment of COVID-19 (Version 6, 2020 Feb 18,http://www.nhc.gov.cn/yzygj/s7653p/202002/8334a8326dd94d329df351d7da8aefc2.shtml), the diagnosis of COVID-19 infection is based on an epidemiological history of contact with COVID-19 patients or travel to endemic areas, combined with laboratory testing including viral RNA detection and radiological findings. COVID-19 patients have similarities of the clinical features with other coronavirus infections. Most patients had fever, cough, myalgia and fatigue, and less often showed symptoms of dyspnoea, haemoptysis and diarrhea [10]. Half of the cases had comorbidities such as diabetic, hypertension and cardiovascular diseases [10]. In our study, we also found some cases that had underlying diseases but without any association with the disease outcome. Age is a risk factor for a more severe disease outcome, because of the generally inferior function of the immune system among older people. Based on the guideline, the patients who have normal body temperature for more than three days, mitigation of respiratory symptoms, improvement of radiological evidences for lesions, and had been tested negative for specific SARS-CoV-2 RT-PCR at least twice, can be discharged from the hospital. The average hospitalized time was 17 days (IQR 11.5 – 21.5), and even as long as 23 days for the patients that were released from our study [5], suggesting the time for SARS-CoV-2 clearance is also around 17 days or longer. This was also been observed in an earlier study where the shedding of SARS-CoV-2 in saliva continued up to 11 days [17].

Until now,there is no effective antivirals available against SARS-CoV-2. To eliminate the virus an effective immunological response with a minimum of immunopathological effects is required [18]. The antigen presentation requires inhibitory alveolar macrophages and dendritic cells (DCs). Successful stimulation can produce viral specific CD4 T cells, which are involved in the development of a specific humoral response, including neutralizing antibodies that can block the viral entry to the cells [19]. CD8+ cytotoxic T cells are required for recognition and killing of the infected cells, further playing a crucial role in SARS-CoV clearance [20, 21]. However, in the biopsy samples from the patients who died of COVID-19, histological examination showed pulmonary oedema with hyaline membrane formation, and interstitial mononuclear inflammatory infiltrates in the lung tissue, suggesting that acute respiratory failure was the cause of death, underlying mechanism that an over-activated immunity injured the lung tissue [12]. Among SARS patients, high expression of dysregulated chemokines and cytokines were found in blood and lungs [22], due to the reason of aberrant immune response caused by the viruses [13, 14]. The delayed or suboptimal immune responses would make chances for SARS-CoV and MERS-CoV to escape from the immune responses, and survive and replicate in the host cells, leading a further delay of the viral clearance [15, 23]. In our study, COVID-19 patients at older ages needed a longer time for the recovery, likely due to the fact that aging is associated with a set of functional and structural alterations in the immune system [24]. Immune-modulators, as corticosteroids, have been empirically used in both SARS and COVID-19 patients [10, 25]. Since corticosteroids might have a decline effect of circulating specific B and T cell subsets [26], the usage on COVID-19 patients should be carefully evaluated before considered [12, 27].

A rapid and generalized lymphopenia has been observed as a prominent part in SARS-CoV and MERS infections [28]. Lymphopenia has also been observed in other viral infections such as measles virus and avian influenza virus, swine foot-and-mouth disease virus, respiratory syncytial virus, and HIV [29-33]. The possible reasons would be lymphocyte sequestration in the lung tissues, or immune-mediate lymphocyte destruction. In MERS infections, the function of bone marrow or thymus suppressed, and furthermore, T cells underwent apoptosis [34-36]. In COVID-19 patients, peripheral CD4 and CD8 T cell counts were found decreased but, they were also hyperactivated, which could result in severe immune injuries [12].

In the present study, we followed the current guidelines and used a specific SARS-CoV-2 realtime RT-PCR for viral detection. Surprisingly, one lymphopenia patient (37y, male, chronic liver disease) was found negative by the realtime RT-PCR and his symptoms had been improved but he is still in hospital. Until now, specific viral RNA detection based on realtime RT-PCR is commonly used in the clinical diagnosis. The usage of realtime RT-PCR is versatile in the detection of SARS-CoV-2 in the respiratory tract specimen or throat swab, but we must keep in mind that those methods always are followed by the probability of false positive/negative. Besides, around 14% of the discharged COVID-19 patients were tested as positive again on follow up samples https://www.caixinglobal.com/2020-02-26/14-of-recovered-covid-19-352patients-in-guangdong-tested-positive-again-101520415.html, showing the recurrence of SARS-CoV-2. This imply that viral detection cannot be used as a single criterion for COVID-19 diagnosis. As a highly contiguous disease with a fatality of approximately 2.3% and up to 49.0% in critical cases [7], every hospitalized COVID-19 patient must be evaluated carefully before discharged. The peripheral leukomonocytes counts could provide more information in the evaluation of COVID-19 patients.

In conclusion, we retrospectively analyzed 25 COVID-19 cases and found that restoration of peripheral leukomonocytes could help the clearance of SARS-CoV-2. We propose here that leukomonocytes counts may serve as a valid prognosis of immune reconstitution for recovery of COVID-19 infection and that can update the current COVID-19 diagnosis guideline.

## Data Availability

All data referred to in the manuscript are availability.

## Acknowledgement

An especially strong appreciation goes to all the clinical physicians, health caregivers, clinical laboratory personals, epidemiologists, and researchers who have been fighting against the SARS-CoV-2 epidemic. We also acknowledge the supports from both nationwide and international resources.

## Reference

1. Li Q, Guan X, Wu P, et al. Early Transmission Dynamics in Wuhan, China, of Novel Coronavirus-Infected Pneumonia. N Engl J Med 2020.

2. Naming the coronavirus disease (COVID-2019) and the virus that causes it. Available at: https://www.who.int/emergencies/diseases/novel-coronavirus-2019/technical-guidance/naming-the-coronavirus-disease-(covid-2019)-and-the-virus-that-causes-it.

3. Available at: https://promedmail.org/.

4. Pan Y, Guan H, Zhou S, et al. Initial CT findings and temporal changes in patients with the novel coronavirus pneumonia (2019-nCoV): a study of 63 patients in Wuhan, China. Eur Radiol 2020.

5. Wang D, Hu B, Hu C, et al. Clinical Characteristics of 138 Hospitalized Patients With 2019 Novel Coronavirus-Infected Pneumonia in Wuhan, China. JAMA 2020.

6. Zhu N, Zhang D, Wang W, et al. A Novel Coronavirus from Patients with Pneumonia in China, 2019. N Engl J Med 2020; 382(8): 727–33.

7. Wu Z, McGoogan JM. Characteristics of and Important Lessons From the Coronavirus Disease 2019 (COVID-19) Outbreak in China: Summary of a Report of 72314 Cases From the Chinese Center for Disease Control and Prevention. JAMA 2020.

8. Bai Y, Yao L, Wei T, et al. Presumed Asymptomatic Carrier Transmission of COVID-19. JAMA 2020.

9. Zhou P, Yang XL, Wang XG, et al. A pneumonia outbreak associated with a new coronavirus of probable bat origin. Nature 2020.

10. Huang C, Wang Y, Li X, et al. Clinical features of patients infected with 2019 novel coronavirus in Wuhan, China. Lancet 2020; 395(10223): 497–506.

11. Chan JF, Yuan S, Kok KH, et al. A familial cluster of pneumonia associated with the 2019 novel coronavirus indicating person-to-person transmission: a study of a family cluster. Lancet 2020; 395(10223): 514–23.

12. Xu Z, Shi L, Wang Y, et al. Pathological findings of COVID-19 associated with acute respiratory distress syndrome. Lancet Respir Med 2020.

13. Wong RS, Wu A, To KF, et al. Haematological manifestations in patients with severe acute respiratory syndrome: retrospective analysis. BMJ 2003; 326(7403): 1358–62.

14. Lau SK, Lau CC, Chan KH, et al. Delayed induction of proinflammatory cytokines and suppression of innate antiviral response by the novel Middle East respiratory syndrome coronavirus: implications for pathogenesis and treatment. J Gen Virol 2013; 94(Pt 12): 2679–90.

15. Liu J, Zheng X, Tong Q, et al. Overlapping and discrete aspects of the pathology and pathogenesis of the emerging human pathogenic coronaviruses SARS-CoV, MERS-CoV, and 2019-nCoV. J Med Virol 2020.

16. Chen H, Guo J, Wang C, et al. Clinical characteristics and intrauterine vertical transmission potential of COVID-19 infection in nine pregnant women: a retrospective review of medical records. The Lancet 2020.

17. To KK, Tsang OT, Chik-Yan Yip C, et al. Consistent detection of 2019 novel coronavirus in saliva. Clin Infect Dis 2020.

18. Newton AH, Cardani A, Braciale TJ. The host immune response in respiratory virus infection: balancing virus clearance and immunopathology. Semin Immunopathol 2016; 38(4): 471–82.

19. Chen J, Lau YF, Lamirande EW, et al. Cellular immune responses to severe acute respiratory syndrome coronavirus (SARS-CoV) infection in senescent BALB/c mice: CD4+ T cells are important in control of SARS-CoV infection. J Virol 2010; 84(3): 1289–301.

20. Janice Oh HL, Ken-En Gan S, Bertoletti A, Tan YJ. Understanding the T cell immune response in SARS coronavirus infection. Emerg Microbes Infect 2012; 1(9): e23.

21. Zhao J, Zhao J, Perlman S. T cell responses are required for protection from clinical disease and for virus clearance in severe acute respiratory syndrome coronavirus-infected mice. J Virol 2010; 84(18): 9318–25.

22. Jiang Y, Xu J, Zhou C, et al. Characterization of cytokine/chemokine profiles of severe acute respiratory syndrome. Am J Respir Crit Care Med 2005; 171(8): 850–7.

23. Min CK, Cheon S, Ha NY, et al. Comparative and kinetic analysis of viral shedding and immunological responses in MERS patients representing a broad spectrum of disease severity. Sci Rep 2016; 6: 25359.

24. Sadighi Akha AA. Aging and the immune system: An overview. J Immunol Methods 2018; 463: 21–6.

25. Stockman LJ, Bellamy R, Garner P. SARS: systematic review of treatment effects. PLoS Med 2006; 3(9): e343.

26. Olnes MJ, Kotliarov Y, Biancotto A, et al. Effects of Systemically Administered Hydrocortisone on the Human Immunome. Sci Rep 2016; 6: 23002.

27. Wu F, Zhao S, Yu B, et al. A new coronavirus associated with human respiratory disease in China. Nature 2020.

28. Li T, Qiu Z, Zhang L, et al. Significant changes of peripheral T lymphocyte subsets in patients with severe acute respiratory syndrome. J Infect Dis 2004; 189(4): 648–51.

29. Okada H, Kobune F, Sato TA, et al. Extensive lymphopenia due to apoptosis of uninfected lymphocytes in acute measles patients. Arch Virol 2000; 145(5): 905–20.

30. Zitzow LA, Rowe T, Morken T, Shieh WJ, Zaki S, Katz JM. Pathogenesis of avian influenza A (H5N1) viruses in ferrets. J Virol 2002; 76(9): 4420–9.

31. Bautista EM, Ferman GS, Golde WT. Induction of lymphopenia and inhibition of T cell function during acute infection of swine with foot and mouth disease virus (FMDV). Vet Immunol Immunopathol 2003; 92(1-2): 61–73.

32. O’Donnell DR, Carrington D. Peripheral blood lymphopenia and neutrophilia in children with severe respiratory syncytial virus disease. Pediatr Pulmonol 2002; 34(2): 128–30.

33. Li CX, Li YY, He LP, et al. The predictive role of CD4(+) cell count and CD4/CD8 ratio in immune reconstitution outcome among HIV/AIDS patients receiving antiretroviral therapy: an eight-year observation in China. BMC Immunol 2019; 20(1): 31.

34. He Z, Zhao C, Dong Q, et al. Effects of severe acute respiratory syndrome (SARS) coronavirus infection on peripheral blood lymphocytes and their subsets. Int J Infect Dis 2005; 9(6): 323–30.

35. Yang Y, Xiong Z, Zhang S, et al. Bcl-xL inhibits T-cell apoptosis induced by expression of SARS coronavirus E protein in the absence of growth factors. Biochem J 2005; 392(Pt 1): 135–43.

36. Mubarak A, Alturaiki W, Hemida MG. Middle East Respiratory Syndrome Coronavirus (MERS-CoV): Infection, Immunological Response, and Vaccine Development. J Immunol Res 2019; 2019: 6491738.

